# Medium-term Prediction of Clinically-relevant Outcomes in First-episode Schizophrenia Patients

**DOI:** 10.64898/2026.03.23.26349083

**Authors:** Eduard Bakstein, Jan Kudelka, Jakub Schneider, Andrea Slovakova, Marketa Fialova, Martin Ihln, Petra Furstova, Jaroslav Hlinka, Filip Spaniel

## Abstract

**BACKGROUND:** Predicting long-term outcomes in first-episode schizophrenia (FES) remains difficult, despite being especially important early in the illness, when timely intervention is most critical. Many potential predictors have been studied, but few are reliable enough to guide early treatment decisions. It also remains unclear how much data from the initial phase of illness is required to improve prognostic accuracy.

**METHODS:** We analysed 68 patients with first-episode schizophrenia (FES) assessed at baseline (V1; mean 0.5 years post-onset, YPO), one-year follow-up (V2; mean 1.2 YPO), and outcome (V3; mean 4.9 YPO). We trained elastic-net models to predict three V3 outcomes—negative symptoms (PANSS Negative factor; Wallwork/Fortgang), global functioning (GAF), and quality of life (WHOQOL-BREF psychological domain)—using either 23 V1 predictors alone or V1 predictors plus V2 data (43 predictors). Performance was evaluated with nested cross-validation on held-out data.

**RESULTS:** Using predictors from the first year (V1+V2), we achieved statistically significant out-of-sample prediction for all three V3 outcomes: PANSS Negative factor (Wallwork/Fortgang) R^2^=0.22— driven mainly by log(DUP), PANSS Negative at V1/V2, and PANSS Disorganized at V2; WHOQOL-BREF Psychological Health R^2^=0.22—driven mainly by WHOQOL Psychological Health at V2 and GAF at V2; and GAF R^2^=0.14—driven mainly by GAF at V2, PANSS Positive at V2, WHOQOL Psychological Health at V2, and hospitalization burden (before V1 and between V1–V2). With baseline-only predictors (V1), only PANSS Negative showed meaningful predictive power (R^2^=0.15); GAF and WHOQOL-BREF did not outperform the intercept-only baseline.

**CONCLUSION:** In FES, long-term functioning (GAF) and quality of life (WHOQOL-BREF) can not be predicted well from first-episode (V1) measures; at least an additional 1 year of follow-up is needed, implying these outcomes are driven by changes after onset that V1 misses. Negative symptoms differ: they are comparatively stable after initial antipsychotic treatment, and duration of untreated psychosis is their strongest predictor beyond baseline severity—consistent with early biology and treatment timing shaping their level and persistence. These contrasting patterns indicate different outcome phenotypes.

## Introduction

The early stage of schizophrenia represents a crucial period characterised by a worsening in cognitive functions (Bora and Pantelis 2015) (Karson et al. 2016) (Bortolato et al. 2015) (Mesholam-Gately et al. 2009), a substantial overall drop in functionality (Salokangas et al., n.d.) (Oomen et al. 2021) (Del Rey-Mejías et al. 2015) and unsatisfactory recovery rate (Huxley et al. 2021). These phenomena unfold in parallel with prominent structural brain changes, suggesting ongoing pathoplastic processes confined to the first years of psychosis (Vita et al. 2012) (Dietsche et al. 2017) (Andreasen et al. 2011).

These findings reinforce the concept of a ‘critical period,’ suggesting that the first 3 to 5 years following diagnosis play a pivotal role in shaping long-term outcomes (Birchwood et al. 1998)(Crumlish et al. 2009) (Wyatt 1991). Additionally, it’s important to note that patients with first-episode psychosis come from diverse backgrounds and have mixed outcomes. Given the available information, we need to stop assuming a one-size-fits-all approach applies to schizophrenia research (Griffiths et al. 2022). Consequently, the early identification of individuals at risk for poor outcomes in schizophrenia may have important clinical implications for tailoring treatment strategies. This could lead to significant advances in the implementation of stratified treatment approaches and potentially halt the progression of dynamic brain changes (Ortiz, Eden, et al. 2017). From this perspective, data-driven outcome prediction closest to the onset of illness gains crucial clinical significance.

An increasing number of studies are being conducted on predicting functional outcomes in individuals experiencing their first episode of schizophrenia (FES). Meta-analyses indicate that sociodemographic, physical, and neuroimaging characteristics have minimal influence on long-term functioning. On the other hand, the duration of untreated psychosis (DUP), cognitive abilities, and simultaneous improvement in both positive and negative symptoms are independently linked to improved functional recovery (Santesteban-Echarri et al. 2017) (Kharawala et al. 2022) (Lee et al. 2022)(Slovakova 2024).

This study aims to determine the clinical, demographic, and functional factors that predict overall functionality and the severity of negative symptoms in people with a first episode of schizophrenia and to create a predictive model that uses early-stage data to forecast these outcomes several years in advance. The findings of this study may aid in identifying patients at high risk, formulating early intervention plans, and monitoring treatment progress in first-episode schizophrenia.

Our research goals were as follows:

1. Predict clinical outcomes in FES patients, including i) the negative symptoms, ii) global functioning, and iii) quality of life five years after illness onset, from demographic and clinical factors obtained during the first year of illness.
2. Determine which of the selected clinical, demographic, and clinical variables are relevant to predicting clinical outcomes after five years.
3. Evaluate the possibilities to predict individual clinical outcomes using data obtained exclusively at the first episode at the onset of the disease

## Methods

### Participants

The study sample comprised 68 patients drawn from the Early-Stage Schizophrenia Outcome (ESO) study. The ESO is a prospective cohort study of patients with first-episode schizophrenia (FES) conducted by the National Institute of Mental Health in the Czech Republic (NIMH) (Spaniel et al. 2016). Patients were recruited through the ESO Patient Enrolment Network, which includes 20 inpatient psychiatric facilities in the Czech Republic. The study is longitudinal, meaning the same examination is repeated at predetermined intervals over several years. Participants undergo a comprehensive examination at three consecutive visits: the first-episode baseline (V1), one year later (V2), and four years later (V3). The study included FES patients who met the following criteria: (1) Age between 18-60 years, (2) Diagnosis of schizophrenia, acute polymorphic psychotic disorder, acute schizophrenia-like psychotic disorder, or schizoaffective disorder as confirmed by a psychiatrist using the International Classification of Diseases-10 (ICD-10) criteria and the Mini International Neuropsychiatric Interview (MINI) (Lecrubier et al. 1997), (3) Experiencing the first episode of psychotic illness, (4) with duration of psychosis less than 24 months, and (5) who are treated with antipsychotic drugs at the time of evaluation.

Excluded were patients with an organic mental disorder, intellectual disability (IQ < 80), a history of seizures, traumatic brain injury with loss of consciousness, intracranial haemorrhage, severe neurological disorder, or substance addiction. The authors ensure that all procedures related to this study comply with the ethical standards set by national and institutional committees on human experimentation and the Declaration of Helsinki of 1975, as updated in 2008. The study was approved by the Ethical Committee of the NIMH, Klecany, Czech Republic (Approval number: 127/17), and all procedures involving human subjects/patients were done with the committee’s approval.

The study sample was selected based on the completion of the third visit (V3) and the completeness of data from the first and second visits (V1 and V2) within selected predictor groups. No missing data were imputed; therefore, only patients with complete data were included in the evaluation.

### Measures of clinical outcome

The primary objective of this analysis is to predict the outcome using three domains, which are clinically significant in both the medium and long term: (1) a functional outcome measure, (2) a measure of the quality of life, and (3) a negative symptom severity measure. The functional outcome (1) was assessed using the Global Assessment of Functioning (GAF) scale, which aims to evaluate an individual’s overall functional adjustment at the visit (American Psychiatric Association and American Psychiatric Association. Task Force on DSM-IV. 2000). Quality of life (2) was measured using the Psychological Health domain from the World Health Organization Quality of Life Brief Questionnaire (WHO-QOL Bref) (Group and THE WHOQOL GROUP 1998). The level of negative symptoms (3) was primarily assessed using the negative factor of the Positive and Negative Syndrome Scale (PANSS) (Kay et al., 1987). At each visit, trained clinicians assessed patients using the PANSS, which we categorised according to Wallwork et al.’s approach (Wallwork et al., 2012) into Positive, Negative, Disorganized, Excited, and Depressed factors. The Negative PANSS factor was selected as the outcome variable because of its documented association with negative symptoms (NS) and long-term outcomes (Milev et al.,(Milev et al. 2005). All target variables were collected at V3, scheduled four years after the first episode (actual mean time from V1 to V3 was 4.9 years in the studied sample)

### Predictor variables

A total of 43 variables from the ESO study sample, collected at V1 and V2, were selected as potential predictors; these included sociodemographic parameters (age, sex, education, etc.) and clinical parameters, including the duration of illness and the duration of untreated psychosis (DUP). For more information on related methodology, see (Slováková et al. 2024), chlorpromazine equivalent dose, number and duration of hospitalizations between V1 and V2, and also the value of the predicted variables at previous time points: psychopathology (PANSS), functional (GAF), and quality of life (WHOQoL). The full list of variables, along with their values across visits, is presented in Table 1 of the Results section. The candidate predictors were selected based on their established relationship with functional outcomes in schizophrenia patients, reported in the literature (Santesteban-Echarri et al. 2017)(Kharawala et al. 2022)(Lee et al. 2022) (Murru and Carpiniello 2018) (Drake et al. 2020). Additional candidate predictors were selected from the ESO study following an exploratory data analysis.

**Table 1:** Overview of the collected dataset. “Y” means “Yes”, “N” means “No”, and “U” means “Unable to determine”.

| Variable |  | M / n | SD / % | Variable |  | M / n | SD / % |
| --- | --- | --- | --- | --- | --- | --- | --- |
| **Measures of outcome** |  |  |  |  |  |  |  |
| Negative Symptoms |  | 1.79 | 0.79 |  |  |  |  |
| Global Assessment of Functioning |  | 79.2 | 13.16 |  |  |  |  |
| WHOQOL Bref Mental Health Domain |  | 14.54 | 2.56 |  |  |  |  |
| **General parameters** |  |  |  |  |  |  |  |
| Age at V1 [years] |  | 27.45 | 6.21 | Interval between V1 and V2 [months] |  | 13.93 | 2.28 |
| Diagnosis | F20.0 | 40 | 59% | Antipsychotic medication (Chlorpromazine equivalent - V1) |  | 375.2 | 204 |
|  | F23.0 | 5 | 7% |  |  |  |  |
|  | F23.1 | 23 | 34% |  |  |  |  |
| Years of education (V1) |  | 15.23 | 3.25 | DUP [months] |  | 2.91 | 4.41 |
| Duration of treatment before V1 [months] |  | 2.93 | 3.98 |  |  |  |  |
| **Clinical parameters** |  | **V1** |  | **V2** |  |  |  |
| Duration of illness (V1) [months] |  | 5.85 | 5.64 | Duration of illness (V2) [months] |  | 19.78 | 6.01 |
| Number of Hospitalizations (V1) |  | 1.35 | 0.77 | Hospitalizations between V1 and V2 |  | 1.15 | 0.74 |
| Days spent hospitalised prior to V1 |  | 44.25 | 43.02 | Days spent hospitalised between V1 and V2 |  | 31.76 | 36.02 |
|  |  |  |  | Days since last hospitalisation (V2) |  | 334.3 | 161.9 |
|  |  |  |  | Hospitalisation status (V2) Currently: |  | 3 | 4% |
|  |  |  |  | In last six months: |  | 10 | 15% |
|  |  |  |  | Not recently: |  | 55 | 81% |
| **Psychopathology** |  | **V1** |  | **V2** |  |  |  |
| Schneider’s sympt. (V1) | Y | 49 | 72% | Schneider’s sympt. (V2) | Y | 29 | 43% |
|  | N | 16 | 24% |  | N | 6 | 9% |
|  | U | 3 | 4% |  | U | 33 | 48% |
| Affective sympt. (V1) | Y | 14 | 21% | Affective sympt. (V2) | Y | 13 | 19% |
|  | N | 45 | 66% |  | N | 23 | 34% |
|  | U | 9 | 13% |  | U | 32 | 47% |
| **PANSS Factors** |  | **V1** |  | **V2** |  |  |  |
| Positive sympt. (V1) |  | 2.02 | 0.78 | Positive sympt. (V2) |  | 1.45 | 0.76 |
| Negative sympt. (V1) |  | 2.42 | 0.89 | Negative sympt. (V2) |  | 2.11 | 0.88 |
| Disorganized sym. (V1) |  | 1.99 | 0.75 | Disorganized sym. (V2) |  | 1.76 | 0.60 |

| Variable | M / n | SD / % | Variable | M / n | SD / % |
| --- | --- | --- | --- | --- | --- |
| Depressed sympt. (V1) | 1.99 | 0.75 | Depressed sympt. (V2) | 1.72 | 0.72 |
| Excited sympt. (V1) | 1.30 | 0.47 | Excited sympt. (V2) | 1.17 | 0.28 |
| QoL and functioning |  |  |  |  |  |
| V1 |  |  | V2 |  |  |
| GAF (V1) | 66.94 | 16.72 | GAF (V2) | 78.79 | 13.80 |
| QOL Physical (V1) | 14.32 | 2.53 | QOL Physical (V2) | 15.26 | 2.26 |
| QOL Psychological (V1) | 13.55 | 2.82 | QOL Psychological (V2) | 14.36 | 2.67 |
| QOL Social relat. (V1) | 13.74 | 2.83 | QOL Social relat. (V2) | 13.71 | 2.92 |
| QOL Enviromental (V1) | 14.58 | 1.98 | QOL Enviromental (V2) | 15.47 | 2.20 |

### Data Analysis

Before building statistical models, we’ve taken several preprocessing and exploratory steps to improve the integrity and homogeneity of our dataset. The exploratory analysis included assessing data distributions and the correlation between variables, which resulted in the following preprocessing and data cleaning steps:

1) All patients with the first episode at an age greater than 40 were removed from our sample as they represented a small minority, differing from the rest of the cohort in multiple demographic and outcome aspects. 2) One patient with schizoaffective disorder was removed from the sample to avoid model overfitting to diagnosis. 3) All patients with a time interval between visit one and visit two that exceeded two years were also excluded from the analysis for consistency. This approach allowed us to focus on a more homogeneous group of patients and minimise any modelling biases arising from outliers or extended time intervals. The data flow diagram illustrating this filtering procedure is shown in Figure 1.

**Figure 1:**
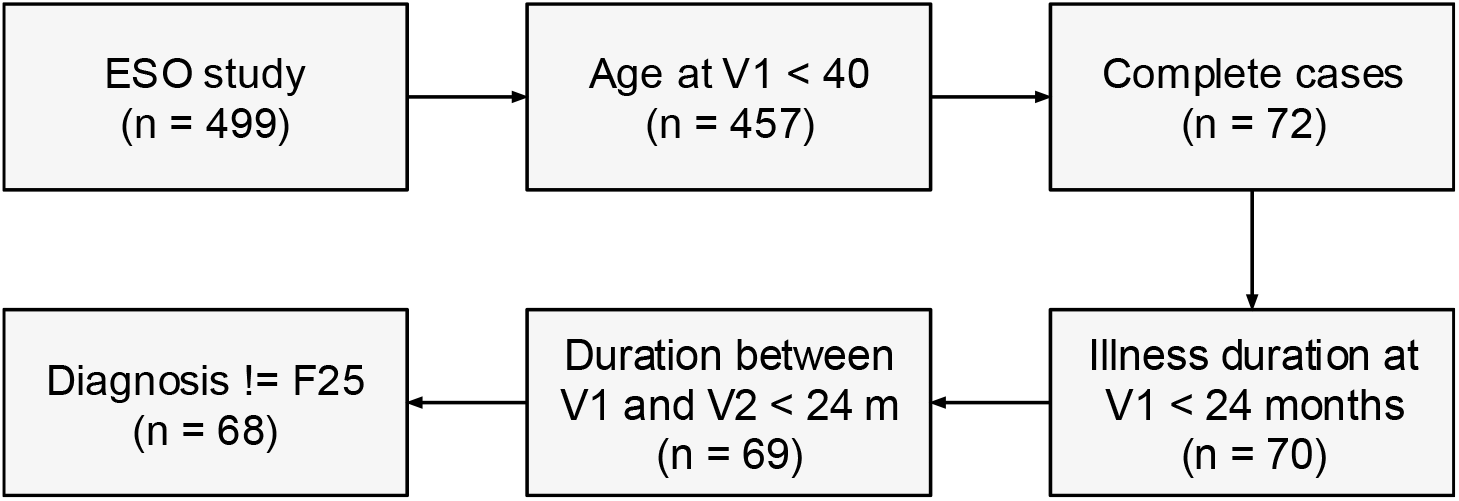
Data flow diagram. The number of observations remaining after each step is displayed.

### Statistical models and validation

In order to evaluate the predictive power of predictors from the first episode vs the first year of disease, we trained two model variants for each outcome variable: i) a model based solely on data from the first episode (V1), and ii) A model combining predictors from the first episode with one year follow-up (V1+V2). To identify the most influential biological factors affecting the outcome, we evaluated the variable importance of individual variables in successful models.

We employed the Elastic-net regression model to address the challenge of a large number of variables (43, respectively, 22) relative to the number of observations (n=68) and to handle multicollinearity, as indicated by moderate correlations between some pairs of variables. The Elastic-net regression combines L1 and L2 regularisation, performs implicit variable selection, and effectively addresses multicollinearity (Zou and Hastie 2005). Elastic-net regression is characterised by two hyperparameters: alpha, the ratio of L1 to L2 regularisation, and lambda, the regularisation constant. During model tuning, values of the lambda parameter were selected using the one-standard-error rule: choosing the largest lambda whose cross-validated error lies within one standard error of its minimum value. Alpha was chosen to minimise the cross-validated error in the nested cross-validation.

The nested cross-validation (Stone 1977) approach was employed to tune model parameters and evaluate models’ performance on unseen data. This approach enables more robust estimation of each model’s performance and proper parameter tuning. Nested cross-validation, an extension of traditional cross-validation, employs a nested structure comprising an outer loop and an inner loop. In the outer loop, the data is divided into a training set and a test set. Within the inner loop, the training set is further subdivided into a training and a validation set. The model is trained on the training set, hyperparameters are fine-tuned on the validation set, and the final model is evaluated on the test set. Specifically, we have employed a 5-fold inner-loop cross-validation nested within each of the 5 outer folds of 5-fold outer-loop cross-validation.

Model performance was evaluated using root mean squared error (RMSE), mean absolute error (MAE), and R-squared (R^2^). Each performance metric was aggregated by averaging across all outer cross-validation folds to estimate model performance on unseen data. Only models achieving lower MAE than the baseline model (an intercept-only variant) on the unseen data were considered successful.

Variable importance was subsequently calculated as the absolute coefficient value divided by the respective variable’s standard deviation on the complete dataset. Data analysis was conducted using the R statistical software, with elastic-net regression models fitted and evaluated using the nestedcv (Comprehensive R Archive Network (CRAN) 2025) and glmnet (Lewis et al. 2023) packages.

## Results

### Dataset description

The summary of the collected dataset variables is presented in Table 1. The data flow diagram, depicting the data filtering process, is shown in Figure 1. Antipsychotic medication at the V2 visit was omitted due to missing values and no apparent relationship with any of the outcome measures. Following exploratory analysis, the logarithm transformation was applied to the DUP.

### Prediction Model Results

The results of the outer cross-validation (i.e., model evaluated on unseen data) for both model variants (using predictors from V1 only /V1/ and using predictors from both V1 and V2 /V1+V2/) for each outcome measure are presented in Table 2. This table also includes variable importance scores and the individual model parameters used for each predicted variable. Predictors of clinical outcome in successful models that exceeded the baseline model on unseen data are depicted in Figure 2.

**Table 2:** Predictive model summary. Each column contains the variable importance scores, performance scores, and parameters for the respective prediction task.

| Predicted outcome: | Negative symptoms<br>(PANSS Negative f.) at V3 |  | Functional outcome<br>(GAF) at V3 |  | Quality of life (WHOQOL)<br>at V3 |  |
| --- | --- | --- | --- | --- | --- | --- |
| Predictors from visits: | V1* | V1 and V2* | V1 | V1 and V2* | V1 | V1 and V2* |
| **Variable importance** |  |  |  |  |  |  |
| Logarithm of DUP | **0.06** | **0.02** | - | - | - | - |
| Negative Symptoms at V1 | **0.26** | **0.12** | - | - | - | - |
| Negative Symptoms at V2 | - | **0.04** | - | - | - | - |
| Disorganized Sympt. at V1 | **0.001** | - | - | - | - | - |
| Disorganized Sympt. at V2 | - | **0.16** | - | - | - | - |
| Excited Sympt. at V1 | **0.02** | - | - | - | - | - |
| Positive Sympt. at V2 | - | - | - | **1.21** | - | - |
| Days spent hospitalized before V1 | - | - | - | **0.35** | - | - |
| Days spent hospitalized before V2 | - | - | - | **0.006** | - | - |
| GAF at V2 | - | - | - | **2.48** | - | **0.07** |
| WHOQOL Psychological health at V2 | - | - | - | **0.57** | - | **0.52** |
| **Performance Metrics** |  |  |  |  |  |  |
| RMSE | **0.72** | **0.69** | **13.46** | **12.14** | **2.63** | **2.32** |
| MAE | **0.61** | **0.57** | **10.38** | **9.52** | **2.03** | **1.79** |
| R-squared | **0.15** | **0.22** | **0.02** | **0.14** | **0.02** | **0.22** |
| **Model Parameters** |  |  |  |  |  |  |
| Intercept | 1.04 | 0.88 | 79.21 | 63.2 | 14.54 | 11.30 |
| alpha | **1.0** | **0.9** | **1** | **0.7** | **1** | **1** |
| lambda | **0.16** | **0.21** | **4.63** | **3.78** | **0.9** | **0.68** |

\* The coefficients and parameters of models whose performance exceeded that of the baseline (intercept-only) model are denoted by an asterisk and printed in boldface.

**Figure 2:**
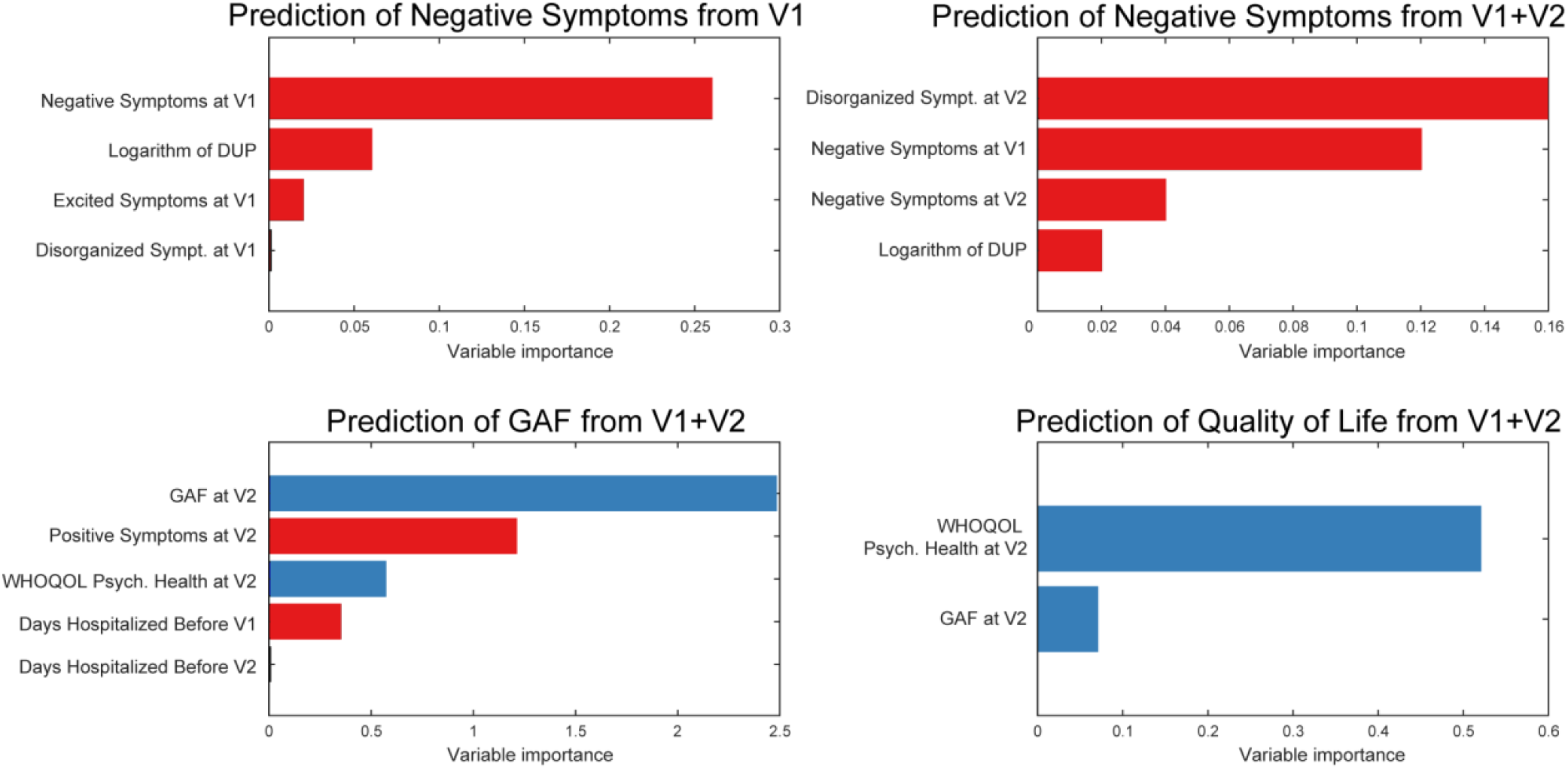
Variable importance scores for the outcome prediction. (A & B) Negative symptoms predicted from values at first episode V1 (A) and in combination with a year follow-up V1+V2 (B). Both model variants achieved significant prediction. For the (C) Global Assessment of Functioning, only the model including predictors from the one-year follow-up (V1+V2) achieved significant predictive performance. For (D), Quality of Life prediction exceeded chance only with the model that included follow-up data (V1+V2). All identified predictors associated with deterioration (increase → deterioration) are marked in red, and those associated with improvement (increase → improvement) are marked in blue.

When predicting negative symptoms at V3 (Fig. 2A and 2B), both model variants (V1 and V1+V2) performed well. Both models included as significant predictors (i) the duration of untreated psychosis (DUP), (ii) the severity of negative symptoms assessed at the first episode visit (V1), and (iii) the presence of disorganised symptoms, utilising the most recent data obtained from the closest available visit.

Concerning the GAF score, only the (V1+V2) model had a better prediction of the GAF score at V3 (Fig. 2C) than the null model (average). The key factors that were associated with the GAF score four years after the onset of symptoms were (i) GAF at V2, (ii) the severity of positive symptoms at V2 (PANSS Wallwork dimension), (iii) the psychological health domain of the WHOQOL-Bref quality of life questionnaire at V2, (iv) the number of hospitalisations before V1 and (v) the number of hospitalisations between V1 and V2. Additional data from a later time point were necessary to make reliable predictions of GAF using the predictor variables analysed.

For Quality of Life (WHOQOL-BREF; see Fig. 2D) prediction, only the V1+V2 model again surpassed the baseline model (average). The only two factors that were found to predict the quality of life four years after baseline were (i) the psychological health domain of the WHOQOL-Bref questionnaire from the one-year follow-up at V2, and (ii) the GAF score at V2.

## Discussion

This study aims to assess the feasibility of predicting medium-term outcomes for patients with first-episode schizophrenia (FES) spectrum disorders five years after the onset of their illness (at V3) from clinical, demographic, and functional predictor variables, collected at first episode visit (V1) or one year after the onset (V2). We employed an elastic-net regression model, which enabled us to handle collinear predictors. To validate the results, we evaluated the model on unseen data using nested cross-validation.

Our primary findings indicate that it is possible to predict negative symptoms severity at V3 solely from predictor values at V1. Adding predictors from V2 substantially improves the model’s predictive performance.

For the two other investigated outcomes - GAF and WHOQOL-Bref - the V1 data were insufficient to build a prediction model that would outperform the baseline (intercept-only) model on unseen patients’ data. Similarly, for both scales, the key predictors were the respective past scale values. In the GAF case, the past value was accompanied by the hospitalization count between V1 and V2. These results suggest that long-term functional impairment in FES is primarily influenced by events occurring at least 1 year after illness onset, making both V1 and V2 time points necessary to predict GAF and quality of life at V3. Also, it should be noted that while the negative symptoms reflect psychopathology directly, the functional outcome scales are affected by a much broader range of socio-economic and environmental factors, and are therefore less directly connected to the disease progression. Therefore, it is unsurprising that early disease development and severity are required as inputs for successful prediction.

Investigating the results for negative symptom prediction further, both V1 and V1+V2 models concur on the importance of (i) the level of negative symptoms (at both V1 and V2 if available), and (ii) the crucial influence of duration of untreated psychosis (DUP), surpassing all other potential predictors, apart from previous symptom severity.

First, these findings suggest that, if present, the disease’s negative symptoms are likely to appear early and persist for a long time, indicating that early manifestation predicts poor future outcomes. Secondly, the prediction of negative symptoms at V3 based on DUP data is noteworthy and in accordance with existing research, which showed a strong association between a prolonged DUP and the increased severity of negative symptoms (Penttilä et al. 2014) (Perkins et al. 2005) (Boonstra et al. 2012). Furthermore, longer DUP is linked to other symptom domains, including an increase in the severity of positive symptoms and a decline in overall functioning (Penttilä et al. 2014) (Perkins et al. 2005) - findings not supported by our study. Studies have suggested increased relapse rates or worse response rates to antipsychotic medications in individuals with longer DUP (Perkins et al. 2005).

The current finding aligns with the hypothesis that negative symptoms (NS) may represent a clinical manifestation of a deteriorating pathological course that seems to be active during the untreated “biologically toxic” stages of psychosis, at least in a subset of cases (Anderson et al. 2014). The frequently reported association between the DUP and NS is notably significant in light of recent findings on the interplay between primary negative symptomatology and inflammation (Dunleavy et al., 2022), as well as the anti-inflammatory effects of antipsychotics (Stamoula et al.,(Stamoula et al. 2022). Overall, DUP’s predictive value underscores the importance of timely initiation of antipsychotic therapy, underscoring its role in preventing future complications.

This study identified several noteworthy determinants of outcome, including minor factors. Specifically, disorganised symptoms were a significant predictor of NS outcomes at V3, as identified by both the V1 and V1+V2 models. Previous research has established the adverse effects of disorganised symptoms in individuals with schizophrenia. It is known that disorganised and negative symptoms are interconnected in schizophrenia, and both are mediators of global functioning in early-onset psychosis (Smelror et al. 2020). Disorganised symptoms in schizophrenia are also associated with poorer overall functionality (Sánchez-Torres et al. 2016) (Liddle 2019) (Ortiz, Gadelha, et al. 2017) (Carrión et al. 2013) (Smelror et al. 2020) (Dominguez et al. 2010).

This study highlights the need for further research to elucidate the potential neurotoxic effects of DUP and the mechanisms underlying structural and functional changes in the early stages of illness. It is essential to comprehend the dynamic processes that occur in one to two years after the onset of an illness, as they show influence on the long-term functional outcomes and quality of life. Furthermore, the study emphasises the need to investigate the impact of symptom persistence and treatment resistance on long-term outcomes. Ultimately, a comprehensive understanding of these dynamic processes may facilitate the development of more efficacious interventions and improved outcomes for individuals with schizophrenia.

### Limitations

The study on patients with first-episode schizophrenia had several limitations that may affect the interpretation of results. Using a sample size of only 68 patients could limit the generalizability of the findings. Additionally, excluding patients with an organic mental disorder, intellectual disability, substance addiction, and onset at age >40 may have further restricted generalizability. The modest sample size was also the reason for using linear models rather than incorporating more complex relationships. However, the highly nonlinear dependencies were addressed by transforming the input variables after exploratory data analysis, as in the case of DUP, which was logarithmically transformed. The study focused on predicting three outcome measures, potentially overlooking other important outcome indicators. Moreover, retrospective data collection may have introduced errors or bias. Finally, selection bias may have occurred because only patients who completed all three visits were included. These limitations should be taken into account when interpreting the study’s results.

## Conclusion

The study’s results suggest that predicting long-term functional outcomes for patients with first-episode schizophrenia spectrum disorders requires gathering data at least one year after the onset of the disease. This highlights the importance of dynamic post-onset events in determining Global Assessment of Functioning and quality of life.

On the contrary, negative symptoms can be predicted using early post-onset data alone, with a longer duration of untreated psychosis being a significant predictor along with symptomatology, warranting further study into the mechanisms behind the duration of untreated psychosis and negative symptoms.

## Data Availability

All data produced in the present study are available upon reasonable request to the authors

## Acknowledgements

This work was supported by the ERDF-Project Brain dynamics, No. CZ.02.01.01/00/22 008/0004643, and by the Ministry of Health of the Czech Republic, grants nos. NU21-08-00432 and NU22-04-00143.

